# Drug name confusion errors in the published literature and the USP Medication Errors Reporting Program, 1965–2004: a reconstructed dataset of 724 confused drug name pairs

**DOI:** 10.64898/2026.10.28.26366036

**Authors:** Bruce L. Lambert, Sengwee Toh

## Abstract

**Background:** Medication errors caused by confusion between similar drug names (look-alike/sound-alike, or LASA, errors) have been reported for decades, but the underlying case reports have never been assembled into a public, machine-readable dataset. Between 2002 and 2005 we compiled such a collection from the published literature and from the United States Pharmacopeia (USP) Medication Errors Reporting (MER) Program. It remained in five unreleased Microsoft Access databases.

**Methods:** In 2026 we exported and reconciled the five databases into case-level and pair-level files. We mapped drug names to RxNorm and to WHO Anatomical Therapeutic Chemical (ATC) classes, computed five orthographic similarity measures, and recorded membership on the FDA and ISMP lists of drug names with recommended Tall Man letters. A limited PubMed search added pairs reported from June 2005 to February 2026. We describe the collection and compare the two sources.

**Results:** The dataset contains 709 literature case records (1965–2004), 753 USP reports (1997–2003), and 17 post-2005 records, covering 724 confused drug name pairs. Only 69 of the 708 original pairs appeared in both sources. Published cases were more severe than voluntary reports: among records with a coded outcome, 15.2% of literature records and 1.2% of USP reports were fatal. Similarity measures discriminated reported pairs from random re-pairings of the same names (area under the ROC curve 0.88–0.92), but 30% of pairs had relatively low measured similarity (BI-SIM below 0.40). Of the pairs with at least three records or at least one fatal-coded record, 122 appear on neither Tall Man list. Automated approximate matching of drug names to RxNorm, used in building the dataset, itself produced name-confusion errors of the kind the dataset records; these were found by manual review and removed.

**Conclusions:** This reconstructed collection is a public historical record of drug name confusion through 2004. It shows how little published and voluntarily reported confusions overlap. It can serve as a benchmark for name-similarity methods and as a starting point for review of confusable names.

## 1. Introduction

Drug name confusion is a recognized source of wrong-drug errors. Names that are similar in spelling and sound are more likely to be confused [4]. Regulators and safety organizations respond with name review before approval and with interventions such as Tall Man (mixed-case) lettering [7]. Research on these problems needs examples of names that have actually been confused, with as much detail about each error as the sources allow.

Two kinds of source exist. Published case reports describe individual errors, often in detail, but favor serious and unusual events. Voluntary reporting programs, such as the USP MER Program, collect many more events, including near misses, but each report is brief. To our knowledge no public case-level collection combines the two.

Toh’s thesis [8] assembled such a collection: case reports extracted systematically from the published literature, combined with USP MER Program reports obtained for research use. The thesis analyzed the data, but the databases themselves were never released. This paper describes the reconstruction of those databases into a public dataset, reports what the reconstructed data show about the two sources, and documents the dataset’s construction and limitations for reuse.

## 2. Methods

### 2.1 Original data collection (2002–2005)

Toh collected the data for a Master’s thesis [8], supervised by Lambert and supported by AHRQ grant R01 HS011609.

#### Literature cases

Case reports were identified by searching PubMed/MEDLINE, International Pharmaceutical Abstracts, Embase, and Google for 1965 to May 2005, using combinations of terms for drug name confusion, medication error, look-alike, and sound-alike. Each article was read in full and its cases coded into an Access database with a standardized coding manual. Coded variables included the two names, which drug was intended and which was given, setting, the roles of the people who made and discovered the error, patient demographics, the NCC MERP outcome category [9], and the reference. A case that involved more than two names was coded once for each confused pair, under a shared case number.

Two additional raters independently coded a random subsample. The variables central to this dataset (the two drug names and the direction of the error) met the preset thresholds of more than 80% agreement and Cohen’s kappa above 0.70, as did the patient and case-report descriptors. Thirteen of the 32 coded variables fell below those thresholds, mostly the more judgment-dependent error and product fields (type of error, outcome category, person who made the error, setting, and origin of error) [8]. All discrepancies were resolved by consensus before the database was finalized, so the released values are reconciled codings, not independent ones.

#### USP MER Program reports

The USP/ISMP MER Program is a voluntary system through which health professionals report actual and potential medication errors [1, 8]. In October 2002 Lambert asked USP in writing for research access to name-confusion reports. USP agreed [2] and delivered an extract in 2004. The reports describe error events and contain no patient identifiers.

USP supplied the extract already restricted to name-confusion reports; we applied no further eligibility filter. The file had 4,173 rows covering 753 reports, usually one row per drug, with extra rows where a report listed several strengths or dosage forms. We kept the first row for each combination of report and drug position (1,506 rows) and combined the two drug positions into one row per report, giving 753 reports and 424 unique pairs. Alternate strengths and dosage forms listed within a report are therefore not represented at the pair level (347 of 1,506 drug positions listed more than one product row).

The reports span ten USP error types: brand names look alike (385), generic names look alike (91), nomenclature (90), brand names sound alike (73), suffix problems (35), brand and generic names look alike (35), generic names sound alike (14), brand and generic names sound alike (13), name similar to a former name (10), and prefix problems (7). The nomenclature, suffix, prefix, and former-name types include errors in which a single name was ambiguous across products. These produce ten pairs with the same name in both positions (e.g., Prelone syrup released in a second strength without a distinguishing name).

### 2.2 Reconstruction and reconciliation (2026)

The five Access databases overlapped. We exported all tables to CSV and reconciled them as follows.

1. **DrugNameConfusions**, the primary database: 695 verified literature case records in 22 related tables. Foreign keys were resolved to readable values.
2. **Literature-based Case Reports**, an earlier thesis snapshot of 584 cases, all contained in DrugNameConfusions. Kept for provenance only.
3. **Lambert working set**: 95 cases compiled by Lambert from ISMP Medication Safety Alerts (2002– 2004), medication error publications by Cohen and Davis (1991–1994), Teplitsky (1994), and other sources. Matching on drug pair and case details found 80 already in DrugNameConfusions; of the 15 others, one was a duplicate on manual review, so 14 records were added (695 + 14 = 709).
4. **USP Case Reports**: the 753 reports described above.
5. **Canonical pair list**: the thesis master list of 367 pairs, expanded to 708 by taking the union with all pairs in the literature (353) and USP (424) case files.

Pair keys were built from alphabetically sorted, lowercased, whitespace-trimmed names so that formatting differences would not create duplicates. Pairs are unordered.

#### Post-2005 search

In February 2026 we searched PubMed for June 2005 to February 2026 with the query (“drug name” OR “medication name” OR “look-alike” OR “sound-alike” OR “LASA”) AND (“confusion” OR “error” OR “mix-up” OR “wrong drug”) AND (“case report” OR “case study” OR “incident”). Of 267 results, title and abstract screening rated 29 as likely and 78 as possible case reports. The likely reports yielded 16 new pairs (17 records), none already in the dataset. This search was a limited addition, not an update of the original review.

### 2.3 Enrichment

All enrichment used Python scripts, which are included in the deposit. API responses are cached so that reruns do not depend on the current state of the APIs. Enrichment was run on the 708 original pairs; the 16 post-2005 pairs were added afterward.

#### RxNorm

Each of the 988 distinct names in the original pairs was looked up with the NLM RxNorm API [3]: exact match first, then approximate match (accepted with a score of at least 5.0), then a match after stripping common salt suffixes. For brand names, the generic ingredient was resolved through RxNorm’s ingredient relations. All accepted matches were then reviewed by hand (Section 2.5).

#### ATC classification

ATC codes for RxNorm-mapped drugs were obtained from the NLM RxClass API [6] at Level 4. For each pair we recorded whether the two drugs share a class at Levels 1–4.

#### Orthographic similarity

Five measures were computed for all pairs:

- **BI-SIM**: Kondrak’s n-gram similarity [10], which aligns overlapping bigrams by dynamic programming with partial credit for character matches, normalized by the length of the longer string. Bigram-based measures have been used to study drug name confusion since the 1990s [4]. **Bigram Dice coefficient**: 2 × shared bigrams / (bigrams in name 1 + bigrams in name 2).
- **Normalized edit similarity**: 1 − (Levenshtein distance / length of the longer name).
- **Jaro-Winkler similarity** [5], which gives extra weight to a shared prefix.
- **Trigram Dice coefficient**: the Dice coefficient on character trigrams.
- **Tall Man lists**. Each pair was checked against the lists published together by ISMP [7]: the FDA-approved list of established names with Tall Man letters (24 pairs) and the combined list of FDA and ISMP-recommended pairs (99 pairs, which includes the FDA pairs).

### 2.4 Analysis

We report counts and percentages. Severity distributions are given separately for each source, excluding literature records whose outcome could not be assigned an NCC MERP category. Because a literature case involving several names is coded once per pair, case and death columns count records, not distinct incidents or patients. We also give the number of distinct case numbers.

To check whether the similarity measures separate confused pairs from arbitrary ones, we built 2,124 control pairs (three per original pair) by randomly re-pairing names from the 708 original pairs. Self-pairs and pairs that appear in the dataset were excluded. We report the area under the ROC curve (AUC) for each measure. This benchmark compares reported pairs with random pairs drawn from the same vocabulary. It does not estimate how well a measure would flag confusable names prospectively; random pairs are likely easier to separate than the plausible alternatives a screening method must handle.

### 2.5 Technical validation

- Row counts were checked against the totals recorded in the thesis databases (695 literature cases, 4,173 USP rows, 367 pairs), and ten random rows were inspected after each transformation.
- In July 2026 the dataset was audited twice: first by OpenAI Codex, then by a separate Claude session that checked each of Codex’s claims against the data files. Six defects were confirmed and corrected, and the reported figures were re-derived from the released files (37 of 37 checks passed).
- In October 2026 a second independent review (OpenAI Codex) recomputed every number in the draft of this paper from the released files, and all five similarity scores for all 724 pairs; all reproduced. It found that pair names differing in capitalization from the drug dictionary had lost their RxNorm and ATC fields at the pair level (24 pairs). These were filled from the dictionary. It also identified the record-versus-incident distinction described in Section 2.4, a personal name in one literature reference (now withheld), and several inaccurate codebook descriptions, all corrected.
- **Drug name confusion in the matching step**. A manual review of every accepted RxNorm match then found 24 that resolved to a different product: 22 of the 52 approximate matches and 2 of the 566 exact matches. Several were look-alike errors of exactly the kind this dataset records. Nolvadex (tamoxifen) was matched to Nolvadent, Lidex (fluocinonide) to Bidex, Primacor (milrinone) to Primacur, and IL-II (interleukin-2) to angiotensin II. In some cases a name was matched to the very drug it had been confused with in the source report: Ultracef (cefadroxil) to Ultracet, E-Vista (hydroxyzine) to Evista, and camphorated tincture of opium to opium tincture. These errors had passed both audits, which confirmed that the figures reproduced but not that each match identified the right drug. The 24 names are now treated as unmatched, with the reason for each recorded in the deposited correction script, and all figures below reflect the correction.
- The released USP file was compared column by column with the internal file; it differs only by two withheld columns (practitioners’ free-text narratives and USP accession numbers), and no remaining text field exceeds 100 characters.

## 3. Results

### 3.1 Composition

The dataset contains 1,479 case records: 709 from the literature, 753 USP reports, and 17 from the post-2005 search (Table 1). The 709 literature records come from 679 distinct case numbers. They cover 724 confused pairs and 1,036 distinct names (1,016 ignoring case).

**Table 1.** Composition of the dataset.

| Measure | Value |
| --- | --- |
| Case records, total | 1,479 |
| Literature records, 1965–2004 (distinct case numbers) | 709 (679) |
| USP MER Program reports, 1997–2003 | 753 |
| Post-2005 literature records (pairs) | 17 (16) |
| Confused pairs | 724 |
| Pairs in literature only | 284 |
| Pairs in USP reports only | 355 |
| Pairs in both sources | 69 |
| Pairs with a single record | 504 |
| Pairs with five or more records | 56 |
| Distinct drug names (case-sensitive) | 1,036 |
| Names mapped to RxNorm (of 988 in the original pairs) | 594 (60.1%) |
| Names with an ATC class (of 988) | 379 (38.4%) |

The two sources overlapped little. Of the 708 original pairs, 284 appeared only in the literature, 355 only in USP reports, and 69 in both. Most pairs were reported once; 56 had five or more records. Table 2 lists the most frequently reported pairs.

**Table 2.** Pairs with the most case records.

| Pair | Records | Literature | USP | Fatal-coded |
| --- | --- | --- | --- | --- |
| Celebrex / Celexa | 45 | 7 | 38 | 0 |
| Celebrex / Cerebyx | 41 | 7 | 34 | 0 |
| Seroquel / Serzone | 33 | 15 | 18 | 3 |
| Depakote / Depakote ER | 24 | 3 | 21 | 0 |
| chlorpromazine / chlorpropamide | 18 | 16 | 2 | 6 |
| Zyprexa / Zyrtec | 17 | 8 | 9 | 0 |
| Navane / Norvasc | 17 | 12 | 5 | 0 |
| Zantac / Zyrtec | 16 | 9 | 7 | 0 |

### 3.2 Severity

Outcomes differed sharply between the sources (Table 3). Of the 409 literature records with a coded outcome, 60.9% involved harm (categories E–I) and 15.2% were fatal. Of the 753 USP reports, 12.5% involved harm and 1.2% were fatal, while 32.0% were category A (circumstances with capacity to cause error). For 300 literature records no outcome category could be coded.

**Table 3.**
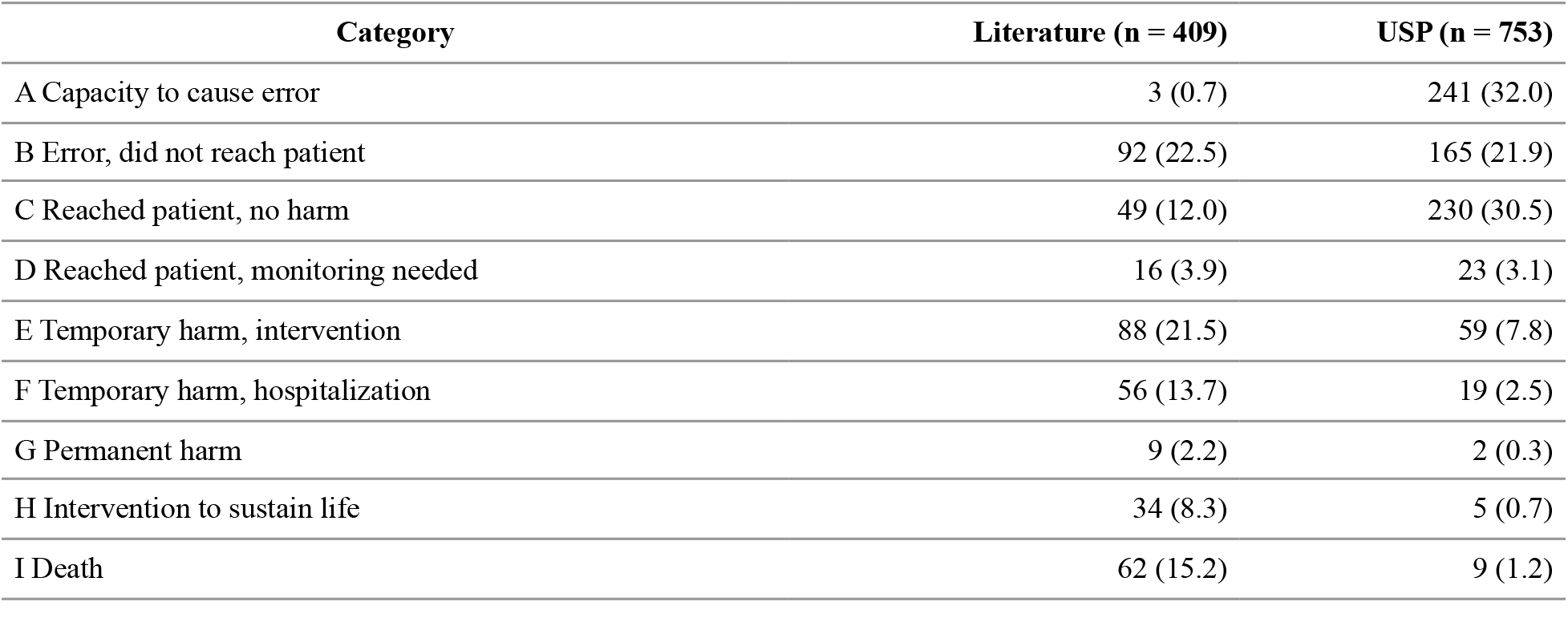
NCC MERP outcome category by source. Number (%) of records with a coded category.

| Category | Literature (n = 409) | USP (n = 753) |
| --- | --- | --- |
| A Capacity to cause error | 3 (0.7) | 241 (32.0) |
| B Error, did not reach patient | 92 (22.5) | 165 (21.9) |
| C Reached patient, no harm | 49 (12.0) | 230 (30.5) |
| D Reached patient, monitoring needed | 16 (3.9) | 23 (3.1) |
| E Temporary harm, intervention | 88 (21.5) | 59 (7.8) |
| F Temporary harm, hospitalization | 56 (13.7) | 19 (2.5) |
| G Permanent harm | 9 (2.2) | 2 (0.3) |
| H Intervention to sustain life | 34 (8.3) | 5 (0.7) |
| I Death | 62 (15.2) | 9 (1.2) |

There were 72 fatal-coded records: 62 in the literature, 9 in USP reports, and 1 from the post-2005 search. The 62 literature records come from 54 distinct case numbers. Thirty-nine pairs have at least one fatal-coded record; the pairs with the most are chlorpromazine/chlorpropamide (6), Isordil/Plendil (5), amiodarone/amrinone (4), and Lasix/Losec (4).

### 3.3 Orthographic similarity

Mean BI-SIM was 0.500 (SD 0.199) for confused pairs (0.493 excluding the ten same-name pairs) and 0.192 for control pairs. All five measures discriminated the 708 original pairs from controls: AUC 0.920 for BI-SIM, 0.917 for trigram Dice, 0.912 for normalized edit similarity, 0.909 for Jaro-Winkler, and 0.878 for bigram Dice. Pairs found in both sources had higher mean BI-SIM (0.559) than pairs found only in the literature (0.532) or only in USP reports (0.470).

Similarity was far from uniform: 221 of the 724 pairs (30%) had BI-SIM below 0.40. These are reported confusions with relatively low measured orthographic similarity; the data do not show why they were confused.

### 3.4 Therapeutic class and ingredient

Both drugs had an ATC class in 188 pairs. Of these, 51.6% shared a Level 1 group (anatomical main group), 30.9% Level 2, 26.1% Level 3, and 21.8% Level 4. Twenty-six pairs resolved to the same ingredient in RxNorm, that is, different products of the same drug (e.g., Depo-Medrol and Solu-Medrol).

### 3.5 Tall Man lists

Nine pairs are on the FDA Tall Man list, and 33 on the combined FDA and ISMP list, including those nine. Pairs on the combined list had more records on average than other pairs (4.3 vs 1.9). Another 122 pairs had at least three records or at least one fatal-coded record but appear on neither list. They include 34 of the 39 pairs with a fatal-coded record, among them Isordil/Plendil, amiodarone/amrinone, and Lasix/Losec.

## 4. Discussion

This collection documents drug name confusions published in English-language literature through 2004 or reported to USP in 1997–2003. Reconstructing it shows three things.

First, the two sources capture largely different confusions. Only 69 of 708 pairs appear in both, and the published cases are far more severe: 15.2% of coded literature records were fatal, against 1.2% of USP reports. Published reports favor serious and unusual outcomes, while voluntary reports capture more near misses. Neither source alone describes the range of name confusion, and neither supports estimates of how often errors occur.

Second, orthographic similarity separates reported pairs from random ones, as earlier work found [4], but many reported confusions involve names with low measured similarity. A screening method based on spelling alone would miss these pairs. The causes of these errors, such as sound, context, packaging, or handwriting, cannot be determined from the records here.

Third, regulatory Tall Man lists cover a small part of the reported confusions. Thirty-four of the 39 pairs with a fatal-coded record are on neither the FDA nor the combined FDA and ISMP list. Some of these names were later withdrawn or changed; amrinone, for example, was renamed inamrinone because of its confusion with amiodarone [1]. List inclusion also reflects considerations other than report counts, so this is not by itself evidence that the lists are incomplete. It does identify pairs that merit review.

The construction of the dataset itself illustrates the problem. Approximate string matching, a reasonable way to tolerate misspellings, mapped Nolvadex to Nolvadent and Ultracef to Ultracet. A similarity score above a threshold is not evidence that two names refer to the same drug, for software any more than for a pharmacist, and checks that only confirm that results reproduce will not catch it.

### Limitations

The collection is a historical record. It reflects what was published through 2004 and reported to USP in 1997–2003, and it makes no claim to be current. The 16 post-2005 pairs come from a single PubMed search with title and abstract screening only; IPA, Embase, ISMP alerts, and FDA sources were not searched. Well-known later confusions are therefore missing; for example, Brilinta (ticagrelor) and Brintellix (vortioxetine), the subject of a 2015 FDA safety communication and a later Sentinel analysis [11], are not included. The literature is restricted to English-language publications.

Case records are not independent. A case involving several names is coded once per pair, and some case reports describe several incidents. Record counts should not be read as counts of incidents or patients, and frequency comparisons across pairs should account for this dependence. Thirteen of the 32 literature coding variables did not reach the preset reliability thresholds (Section 2.1).

RxNorm coverage is 60.1% and ATC coverage 38.4%. Many unmatched names appear to be older brands, misspellings, or products not marketed in the United States. Each brand was resolved to a single ingredient, so combination products carry only one of their ingredients (Percocet, for example, is recorded as acetaminophen), and ATC classes include classes that RxClass assigns to an ingredient through combination products. The ingredient and class-sharing fields are therefore approximate. RxNorm describes current products, and a brand name reused for a reformulated product may map to the wrong ingredient for the era of a report; we found and removed one such case (Kaopectate) but cannot rule out others. The enrichment and ROC analysis cover the 708 original pairs; for the 16 post-2005 pairs, ATC sharing below Level 1 was not computed.

The counts differ from those in the thesis, which reports 1,675 screened articles, 777 case reports, and 370 pairs. This archive has 1,664 bibliographic records, 709 literature case records, and 367 pairs in the thesis master list. The differences come from the thesis-era restriction to verified cases and late deduplication; the original case-by-case exclusion decisions could not be recovered.

### Reuse

Possible uses include benchmarking name-similarity and name-screening methods against historically confused pairs, studying confused pairs with low orthographic similarity, comparing published and voluntarily reported errors, and identifying candidate pairs for Tall Man lettering or other interventions.

## Data Availability

All data produced in the present study are available online at Zenodo (doi:10.5281/zenodo.23508861): case-level and pair-level data files, the codebook, the processing scripts, and the original 2005 thesis. Data and documentation are released under CC BY 4.0 and code under the MIT License.

https://doi.org/10.5281/zenodo.23508861

## 5. Data records and availability

The dataset is deposited at Zenodo (doi:10.5281/zenodo.23508861) with the files in Table 4. Data files and documentation are released under CC BY 4.0 and code under the MIT License; the thesis is included with its author’s permission. The scripts document how the files were produced; two read regulatory-list and prescribing-frequency files from the authors’ wider project directory and will not run on their own, so the deposited CSV files are the authoritative outputs.

**Table 4.** Files in the deposit.

| File | Rows | Columns | Description |
| --- | --- | --- | --- |
| master_pairs.csv | 724 | 48 | One row per confused pair: names, record counts, severity, similarity scores, RxNorm identifiers, ATC codes, Tall Man list membership. |
| master_drugs.csv | 988 | 13 | One row per drug name in the original 708 pairs: RxNorm mapping and ATC classification. |
| master_codebook.txt | – | – | Column definitions for both master files and notes on interpretation. |
| literature_cases_clean.csv | 709 | 22 | One row per literature case record: pair, direction, number of incidents, outcome, staff roles, setting, patient sex and age where reported, reference. |
| usp_cases_redacted.csv | 753 | 32 | One row per USP report, with narratives and accession numbers removed: pair, USP error type, outcome, setting, staff roles, product attributes, prescribing-frequency estimates. |
| Darren_Toh_Thesis.* | – | – | The 2005 thesis (PDF, Word, Markdown), including the bibliography. |
| scripts.zip, cache.zip | – | – | Processing and correction scripts, and cached API responses (zipped folders). |
| AUDIT_REPORT_2026-07.md | – | – | Report of the July 2026 audit. |
| MANIFEST.sha256 | – | – | SHA-256 checksum of every file in the deposit. |

The pair file stores ATC Level 1 codes and labels and Level 4 codes for each drug, with sharing flags at Levels 1–4. Its `Both_In_NM` column records membership on a local institutional (Northwestern Medicine) Tall Man list, not the FDA or ISMP lists.

## 6. Ethics

The original study was reviewed by the Institutional Review Board of the University of Illinois at Chicago and determined not to require review, since it used only published literature and voluntary error reports without patient identifiers. USP provided its data for research use.

## 7. Author contributions

**Bruce L. Lambert:** conceptualization, supervision, data curation, formal analysis, writing (original draft, review and editing), funding acquisition. **Sengwee Toh:** conceptualization, data curation, investigation, methodology, writing (review and editing).

## 8. Funding and acknowledgements

### Funding

Original data collection (2002–2005) was supported by the Agency for Healthcare Research and Quality (AHRQ), grant R01 HS011609. The 2026 archiving work received no specific funding. The funder had no role in this paper or the release of the data.

## Acknowledgements

Diane Cousins, R.Ph., then Vice President of Practitioner Reporting at USP, arranged access to the MER Program data. Jason Tan contributed to the analysis of non-name product attributes during a USP Fellowship.

## 9. Use of generative AI

The 2026 archiving work was carried out with Claude Code (Anthropic; Claude Opus models), working under the direction of the first author. Claude Code wrote the Python code that exported the Access databases, reconciled them, queried the RxNorm and RxClass APIs, computed the similarity measures, and built the master files. It also drafted the codebook, the README, and this paper. OpenAI Codex and separate Claude sessions were used to audit the data and the manuscript (Section 2.5).

The RxNorm matching code written by Claude Code accepted approximate matches that confused one drug name with another (Section 2.5). A later Claude Code session found these errors on manual review, and the first author reviewed the corrections. We report this because it is an instance of the error the dataset documents, and because neither the code’s author nor two audits caught it until each match was checked by hand.

The original data (2002–2005) were collected and coded by people, without AI. Lambert directed the work, reviewed the outputs, and specified substantive corrections, including the distinction between Kondrak’s BI-SIM and the bigram Dice coefficient and the use of reshuffled rather than arbitrary control pairs. Lambert also decided what to release. Every reference was checked against CrossRef, PubMed, or the issuing organization. The authors take full responsibility for the content of the dataset and this paper.

